# Single-nucleus RNA velocity reveals synaptic and cell-cycle dysregulations missed by gene expression in neuropathologic Alzheimer’s disease

**DOI:** 10.1101/2022.10.31.22281759

**Authors:** Quadri Adewale, Ahmed F Khan, David A. Bennett, Yasser Iturria-Medina

## Abstract

**Background:** Typical differential single-nucleus gene expression (snRNA-seq) analyses in Alzheimer’s disease (AD) provide fixed snapshots of cellular alterations, making the accurate detection of temporal cell dynamics challenging.

**Methods:** To characterize the dynamic genetic and cellular differences in AD neuropathology, we apply the novel concept of RNA velocity to the study of single-nucleus RNA from the cortex of 60 subjects with varied levels of AD pathology. RNA velocity captures the rate of change of gene expression by comparing intronic and exonic sequence counts. We performed differential analyses to find the significant genes driving both cell-specific RNA velocity and expression differences in AD, extensively compared these two transcriptomic metrics, and clarified their associations with multiple neuropathologic traits. The results were cross-validated in an independent dataset.

**Results:** Comparison of AD pathology-associated RNA velocity with parallel gene expression differences reveals sets of genes and molecular pathways that underlie the dynamic and static regimes of cell type-specific dysregulations underlying the disease. Differential RNA velocity and its linked progressive neuropathology point to significant dysregulations in synaptic organization and cell development across cell types. In addition, there are accelerated cell changes in AD subjects compared to controls, suggesting that the precocious depletion of precursor cell pools might be associated with neurodegeneration. Finally, we find active molecular drivers of the spatiotemporal alterations in neuropathological AD and discuss implications towards gene- and cell-centric therapeutic strategies.

**Conclusions:** In sum, our results support that the consideration of other less-studied molecular processes (RNA velocity) offers substantial complementary information to the typical analysis of RNA abundance alone.

## Introduction

Recent advances in single-nucleus RNA sequencing (snRNA-seq) have provided an unprecedented ability to disentangle cellular-level transcriptomic alterations and heterogeneity in Alzheimer’s disease (AD) (Mathys et al. 2019; Lau et al. 2020). Early differential expression with neuropathological AD progression have been found to be cell-type dependent while apparent upregulation of genes in the later stages are shared across cell types, suggesting that transcriptional responses to disease are highly driven by cell states (Mathys et al. 2019). Furthermore, cell-specific analysis has revealed the molecular signatures of preferentially affected cell populations, demonstrating that morphology alone cannot sufficiently determine cell type vulnerability in pathologic AD (Leng et al. 2021; Olah et al. 2020; Bergen et al. 2020).

Nevertheless, static snRNA-seq abundance provides only fixed snapshots of cellular states, not revealing temporal dynamics of genes at individual cells (Bergen et al. 2020). The recently proposed rate of change of mRNA, otherwise known as RNA velocity (RNA-vel) (La Manno et al. 2018), provides a novel method to capture temporal dynamics in mRNA abundance by comparing spliced and unspliced mRNA counts. In the initial model, ratio of intronic to exonic sequence counts in constant (steady-state) transcription is obtained, and RNA-vel is estimated as the deviation or residual of this ratio from the expected steady-state ratio (La Manno et al. 2018). As there is ample reason to think that the steady-state assumption is problematic, recent analytical advances have relaxed the assumption of constant transcription rates (Bergen et al. 2020). Positive and negative RNA velocities imply upregulation and downregulation of a gene, respectively. Notably, RNA-vel analysis was used to infer developmental trajectories of healthy cells (Kanton et al. 2019; Lo Giudice et al. 2019) and unravel pathological changes in cancer cells (Couturier et al. 2020). This paradigm shift from descriptive to predictive RNA modelling is offering a deeper understanding of complex cell-level processes in health and disease, with promising implications to improve treatment strategies for multiple disorders.

In the context of the progression of neuropathology, it is unclear whether genes that are differentially expressed across disease states are also the genes driving evolution and vulnerability of diseased cells. Genes with different RNA velocity values might better capture or capture complementary aspects of the time-resolved molecular dysregulations and prodromal differences underlying neurodegeneration. Here, we extend previous single-nucleus RNA (snRNA) analysis in AD in three fundamental ways. First, we use postmortem snRNA-seq data from the prefrontal cortex of subjects with varied levels of AD pathology (N=48) to identify cell-specific dynamic RNA velocity differences associated with neuropathology. Second, we demonstrate that dynamically altered genes in AD pathology, i.e., genes with differential RNA velocities, are qualitatively different from the genes showing differential expression patterns. Third, we reproduce the main observed gene- and cell-specific RNA velocity differences in an independent pathologic AD sample. Overall, our results highlight the critical importance of further considering dynamic single-cell molecular processes underlying AD progression as opposed to only its static cellular RNA mechanisms.

## Methods

### Dataset-1 (Prefrontal cortex)

It includes droplet-based snRNA-seq, neuropathological and clinical data for 48 participants enrolled in the Religious Orders Study (ROS) or the Rush Memory and Aging Project Study (MAP (Bennett et al. 2018). The snRNA-seq data was previously generated from the prefrontal cortex (Brodmann area 10) of autopsied brains as described (Mathys et al. 2019), and it was downloaded from the Accelerating Medicines Partnership Alzheimer’s Disease knowledge portal (AMP-AD; www.synapse.org, ID syn18485175). All subjects underwent postmortem neuropathologic evaluations, generated in previous ROSMAP studies as described in (Bennett et al. 2018) including uniform structured assessment of AD pathology, and other pathologies common in aging and dementia (downloaded from AMP-AD, ID syn3157322; see also *Correlation with neuropathology* subsection below). The 48 subjects (balanced between sexes) comprised 24 with no or low AD-pathology (control group), and 24 with mild to severe AD-pathology (AD group) as determined by β-amyloid burden, neurofibrillary tangles, and cognitive impairment (Mathys et al. 2019). The subjects were matched for age (medians 87.1 [no pathologic AD, N=24] and 86.7 [pathologic AD, N=24]) and years of education (medians of 18 [no pathologic AD] and 19.5 [pathologic AD]). Both ROS and MAP were approved by an Institutional Review Board of Rush University Medical Center. All participants signed an informed consent and Anatomical Gift Act. In addition, they signed a repository consent allowing their data to be shared (related documents and requests for data can be obtained at https://www.radc.rush.edu). The process of isolating the nuclei from the postmortem brain tissues was previously detailed (Mathys et al. 2019). Briefly, the brain tissue was homogenized at very low temperature and incubated. The tissue was then filtered and purified with working solutions. The nuclei were separated through spinning at high speed and counted. The sequencing libraries were constructed with the Chromium Single Cell 3′ Reagent Kits v.2 (10x Genomics) and sequenced with the NextSeq 500/550 High Output v2 kits (150 cycles).

### RNA abundance and cell type identification

Intronic and exonic counts were obtained using kb-python (v0.26.3), a wrapper for kallisto and bustools (Bray et al. 2016; Melsted et al. 2021). First, index file of the human genome was generated from the Ensembl human primary reference genome sequence and gene annotation (GRCh38). Then, spliced and unspliced RNA counts were obtained by filtering barcodes with low UMI counts and mapping reads to the index file. The counting process was performed by sequentially running ‘kb ref’, and ‘kb count’ (with filter flag set) commands.

Next, we acquired a previous quality controlled list of genes, cells and cell types (Mathys et al. 2019). We then looked for shared genes and cells between our filtered counts and the previously reported list. Thus, we had we 65,422 cells with 16,844 transcripts (corresponding to 16,829 unique genes). These cells were then assigned to cell types, based on the reported list, as excitatory neurons, inhibitory neurons, astrocytes, microglia, oligodendrocytes, oligodendrocyte precursor cells, endothelial cells, and pericytes. Endothelial and pericyte cells were subsequently excluded because of their very low counts or absence in some subjects.

### RNA velocity estimation

We used scvelo (v0.2.3) (Bergen et al. 2020) to calculate RNA velocity. First, the cells were pulled together across all subjects, and each cell was normalized by its total size. The normalization was applied to both spliced and unspliced counts. To estimate RNA velocity using the stochastic method, we computed the means and variances of nearest neighbors of cells in principal component analysis (PCA) space. Here, 100 nearest neighbors and 30 PCA were used. Normalization and moments calculation were achieved through ‘pp.normalize_per_cell’ and ‘pp.moments’ commands, respectively. The RNA velocity is then estimated with ‘tl.velocity’ command (setting the mode to ‘stochastic’).

We next sought to validate the estimated RNA velocities by examining the velocity values of the genes driving cell-specific dynamics. We ran a differential velocity Welch’s t-test with the module ‘scv.tl.rank_velocity_genes’ and obtained the top genes (based on t-value) having cell-specific differential velocity expression. We then projected the velocities and expression values of the dynamic genes into t-SNE space to examine their variations across cell types.

### Differential expression and RNA velocity analyses

Cell type-specific gene analysis was performed with Seurat (v4.0.2) and Presto packages in R. We performed differential expression analysis between the control group and the AD group. Each cell was first normalized by its total count over all genes, scaled by 10,000 and log-transformed. Using the ‘FoldChange’ command in Seurat, we performed Wilcoxon rank-sum test to identify differentially expressed genes at log_2_ (fold change) >0.25 or <-0.25. We then used the Presto package (due to its speed) to run 5000 random permutations by randomly reassigning the subjects to either the control or AD group. The U-statistics from the permutations were used to generate null distributions and significance p-values. We identified significant genes after adjusting for multiple testing (q<0.05, FDR-corrected). To compare differential expression with differential velocity, the procedure was repeated on RNA velocities to identify the dynamic genes driving the velocity difference between the control and AD groups.

### Cell speed and residual velocity estimation

First, the speed of a cell was calculated as the length of its velocity vector. Wilcoxon ranked-sum test was then used to compare the speed between the two groups. Next, the RNA velocities of individual cells were used to derive velocity fields in t-SNE space. For each of the groups (control and AD), we linearly interpolated the velocity fields at the t-SNE coordinates where actual cells are missing to ensure equal number of velocity fields for each group. We then subtracted the velocity fields at same pair of coordinates for the two groups and obtained the z-scores of the norms of these differences. The velocity field difference of those coordinates where the z-score>1.96 or <-1.96 are displayed as the residuals between the two groups.

### Correlation with neuropathology

In each cell type and subject, the average RNA velocity across cells was calculated for every gene. The velocities were tested for correlation with four AD neuropathological traits (Bennett et al. 2018): PHF neurofibrillary tangle density (tangles), neuronal neurofibrillary tangle counts (NFT), overall β-amyloid load (β-amyloid), and neuritic plaque counts (NP). The correlations were adjusted for age, sex, and postmortem interval. Significant genes were chosen based on FWER-corrected p-value < 0.001 (Genovese, Lazar, and Nichols 2002).

### Validation in independent dataset (Dataset-2: Dorsolateral prefrontal cortex)

The droplet-based snRNA-seq data was previously generated from the dorsolateral prefrontal cortex of autopsied brains as described (Cain et al. 2020), and it was downloaded from the Accelerating Medicines Partnership Alzheimer’s Disease knowledge portal (AMP-AD; www.synapse.org, ID syn16780177). The subjects include another 12 sex-matched individuals from Orders Study (ROS) or the Rush Memory and Aging Project Study (MAP) (Bennett et al. 2018): 6 subjects are cognitively non-impaired with minimal AD pathology and 6 fulfill diagnoses for both pathologic AD and clinical AD dementia.

As previously described (Cain et al. 2020), the brain grey matter tissue was homogenized and treated with working solution to separate the nuclei. The isolated nuclei were then counted and filtered. The libraries were constructed and sequenced on the 10X Single Cell RNA-Seq Platform using the Chromium Single Cell 3’ Reagent Kits v2. After obtaining the intronic and exonic counts, genes were selected according to the gene list from Dataset-1 and the cells were filtered using the cell list obtained from the metadata of the previous study (Cain et al. 2020). The previously reported cell clusters were used to assign the 79,472 cells to the six cell types under consideration. We calculated the cell-specific RNA velocities for each subject and used Wilcoxon rank-sum test to identify the genes underlying RNA velocity differences between the minimal AD pathology group and the pathologic/AD dementia group.

We assessed the overlap between the significant differential velocity genes in this dataset and Dataset-1. Fisher’s exact test was used to obtain the significance of overlap (p-value<0.01 and odds ratio>1).

### Biological pathway analyses

Biological pathways were identified using EnrichR online tool to query enriched gene ontology (GO) terms (Chen et al., 2013; Kuleshov et al., 2016) from the Gene Ontological Biological Processes 2021. The significant GO terms were selected at an adjusted p-value<0.01 and ranked based on their EnrichR combined scores.

### Comparison between single-cell and single-nucleus RNA velocities (Dataset-3)

We downloaded a previously published dataset from the Gene Expression Omnibus (GEO) (https://www.ncbi.nlm.nih.gov/geo/, GSE135618). The dataset contains matched single cells, fresh nuclei and frozen nuclei obtained from the microglia of two subjects (Gerrits et al. 2020). After preprocessing, we obtained 2,988 cells, 4,892 fresh nuclei, and 4,019 frozen nuclei from one subject; and 3,485 cells, 2,593 fresh nuclei, and 5,527 frozen nuclei from the other subject. For each of the subjects, we estimated the RNA velocities across the three modalities (single cell, fresh nuclei, and frozen nuclei) and performed within-subject comparisons of the velocity estimates.

### Data visualization

We visualized the data using scvelo (0.2.3) (Bergen et al. 2020) in python, and ComplexHeatMap (v.2.6.2) in R (Zuguang, Roland, and Matthias 2016).

## Results

### Data origin and single-cell RNA velocity estimation

Single nucleus RNA-seq data was obtained from the prefrontal cortex of 48 postmortem human brain samples (Mathys et al. 2019) (*Methods, Dataset-1* subsection). Twenty-four of these individuals had no or low β-amyloid burden or other pathologies (control). The remaining twenty-four presented mild to severe AD-pathology (amyloid burden, neurofibrillary tangles, global pathology, and cognitive impairment). After pre-processing, 65,422 snRNA-seq profiles with 16,844 transcripts (corresponding to 16,829 unique genes) were obtained. A predefined cluster list (Mathys et al. 2019) was used to annotate and assign the cells to six different types: excitatory neurons, inhibitory neurons, astrocytes, microglia, oligodendrocytes, and oligodendrocyte progenitor cells (Figure 1A).

**Figure 1.**
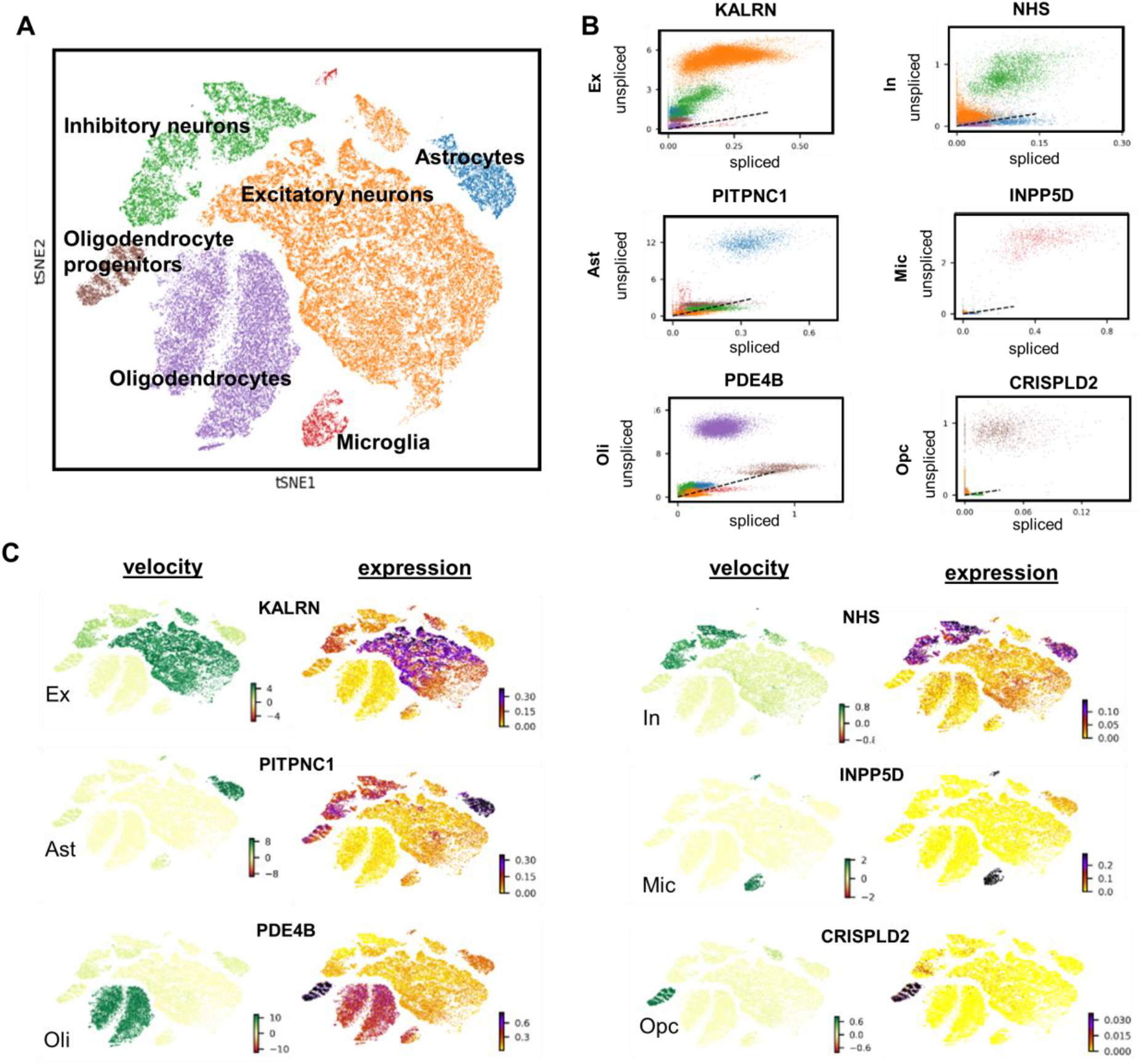
Single-nucleus RNA-seq of the prefrontal cortex of 48 individuals across the Alzheimer’s disease (AD) spectrum. A) t-SNE visualization of clusters are annotated by cell type (excitatory neurons, inhibitory neurons, astrocytes, microglia, oligodendrocytes, and oligodendrocyte progenitor cells). B) Relationship between unspliced and spliced mRNA counts of genes driving differential dynamics of each cell type. The dashed black line represents the estimated steady state ratio of the unspliced to spliced mRNA. RNA velocity is obtained as the residual of the observed intronic to exonic RNA ratio from this steady state line. C) RNA velocity and expression patterns of the dynamic genes. Larger variation in velocity is driven by transcriptional dynamics.

Single-nucleus RNA velocities (snRNA-vel) of all genes across each cell type in the 48 subjects were calculated using a probabilistic model (Bergen et al. 2020) (*Methods, RNA velocity estimation* subsection). This probabilistic (stochastic) method for RNA-vel estimation is preferred over the originally proposed steady-state model (La Manno et al. 2018) since the former largely accounts for cell heterogeneity and differential kinetics, while achieving higher computational efficiency (Bergen et al. 2020). To identify the genes that may help explain the velocity vector fields across the six types, we selected the top genes that show cell-specific differential transcriptional dynamics (*Methods, RNA velocity estimation* subsection). As shown in Figure 1B, the dependency between unspliced and spliced mRNA counts of the genes gives the expected cell-specific velocity values depicted as the residual from the dotted line (representing the constant steady-state transcription). We then projected the expression and velocity values of these top genes to t-SNE space (Figure 1C). We observed more variation and cell-specificity in RNA velocity compared to gene expression, suggesting that the velocity estimations are largely driven by transcriptional dynamics rather than gene expression (Figure 1C). For example, *PDE4B* exhibits an oligodendrocyte-specific dynamics even though its expression is spread across different cell types.

To further demonstrate the suitability of using single nuclei for RNA velocity, we compared both nucleus-derived (snRNA-seq) and cell-derived (scRNA-seq) RNA velocities in the microglia of the same subject. There was a considerable correlation (R = 0.94 – 0.99) between the velocity estimates of the snRNA-seq and scRNA-seq, supporting the validity of snRNA-seq for RNA velocity calculation (Supplementary Figure 1). Interestingly, the variations observed in velocity correlations from any random pair of single cells is comparable to the variations in velocity correlations between any single cell and single nucleus (Supplementary Figure 1C).

### Static vs dynamic genetic-cell modifications underlying AD evolution

We sought to investigate if there are global AD-pathology dependent differences in RNA velocity patterns across cell types. We evaluated the cell-specific differences in RNA velocity between the control and AD-pathology subjects using Wilcoxon rank-sum test. We then compared the results with those obtained for differential gene expression. Across all cell types, we observed lesser genes with differential RNA velocity (612) than those with differential expression (3152) (Figure 2A). The top ranked genes underlying RNA expression and velocity variations are presented in Figure 2B. Furthermore, higher fold changes were observed for RNA velocities compared to gene expression. Due to potential over-representation of long unspliced mRNA transcripts in neurons (Gorin and Pachter 2021), we checked if the differential velocity observed between the two groups may be biased by gene length. We found no correlation (R=0.00081; p-value=0.92) between the U-statistic of Wilcoxon rank-sum test and the length of genes in inhibitory neurons (Supplementary Figure S2).

**Figure 2.**
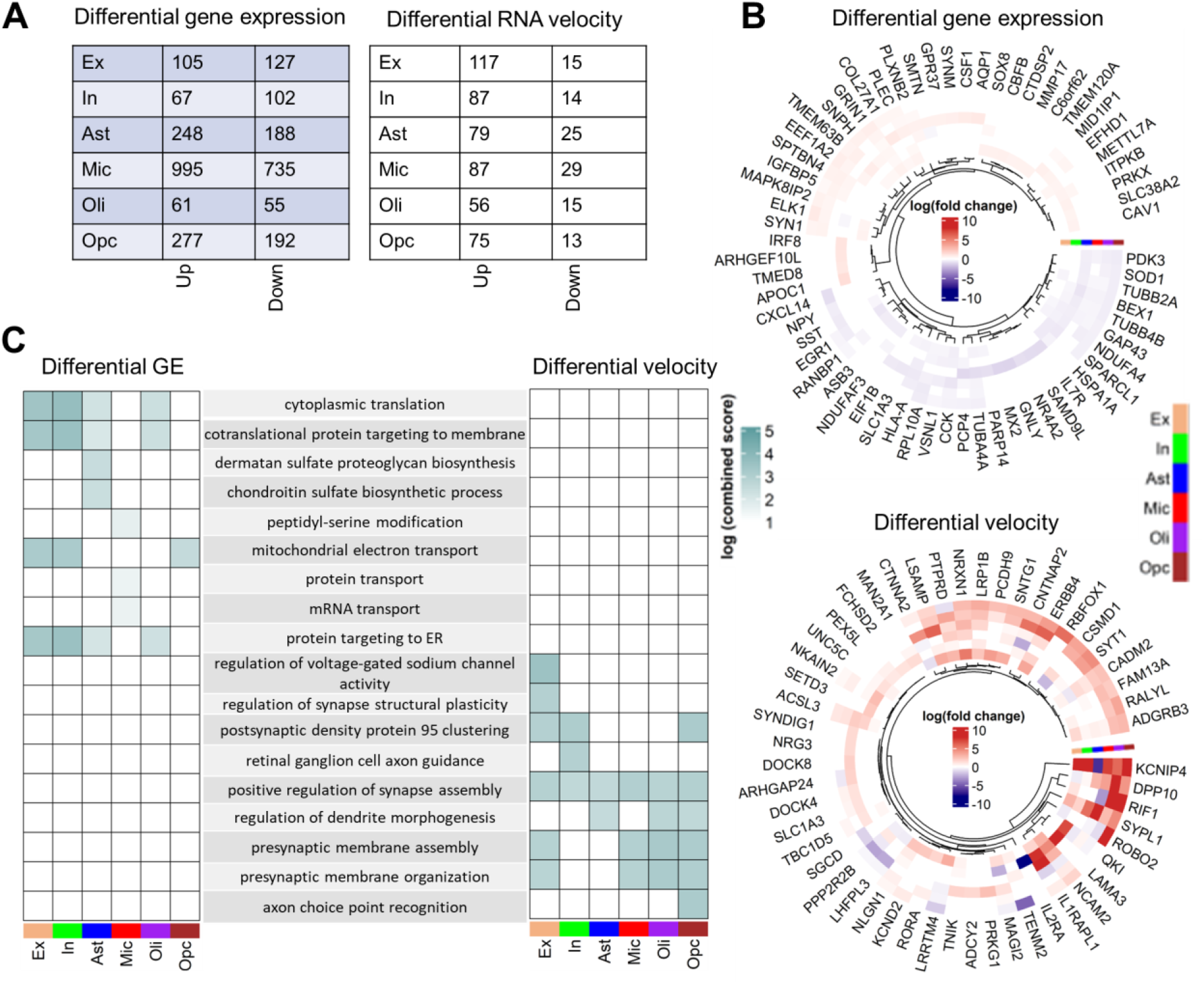
Differences in RNA velocity and gene expression underlying neuropathological AD progression. A) Number of genes with differential expression patterns and velocity between controls (n=24) and AD subjects (n=24) across cell types (two-sided Wilcoxon rank-sum test, permutation-based FDR-corrected q-value < 0.05, log_2_ (mean gene expression or velocity AD/mean gene expression or velocity in control) > 0.25 or < −0.25). B) Cell-type specific changes (log_2_ (fold change)) for the top genes with differential expression (DE) (top) and differential velocity (bottom) between control and AD subjects. C) Comparison of biological pathways associated with differential expression and differential velocity.

Notably, only 63 of the 3152 (2%) differentially expressed genes were also found to exhibit differential velocity, suggesting substantial AD-pathology related differences between these two RNA descriptors. The genes with only snRNA-vel differences relate to cell developmental and synaptic processes such as morphogenesis, axonal guidance, ion channel activity, synapse organization and cell assembly (Figure 2C). Conversely, the genes with only differential expression are majorly associated with mitochondrial activity, ribosomal processes, and protein sorting. This mismatch between snRNA-vel and RNA abundance dysregulations suggests that analyzing RNA velocity provides relevant complementary information about the multifactorial molecular processes associated with neuropathological AD advance compared to differential expression.

### Several RNA-velocity differences underlie AD neuropathological severity

We proceeded to conceptualize the observed differences in RNA velocities between the control and AD groups. We projected the velocity vectors into t-SNE space and evaluated the group difference in velocity fields across all cells. Figure 3A shows the residual velocity fields which account for the difference between the two groups. To further understand the biological implication of the observed residual velocity fields, we calculated the speed of individual cells using the velocities across all genes. Indeed, we found a higher speed in the AD groups compared to controls, suggesting that accelerated cell changes are associated with AD neuropathology (Figure 3B).

**Figure 3.**
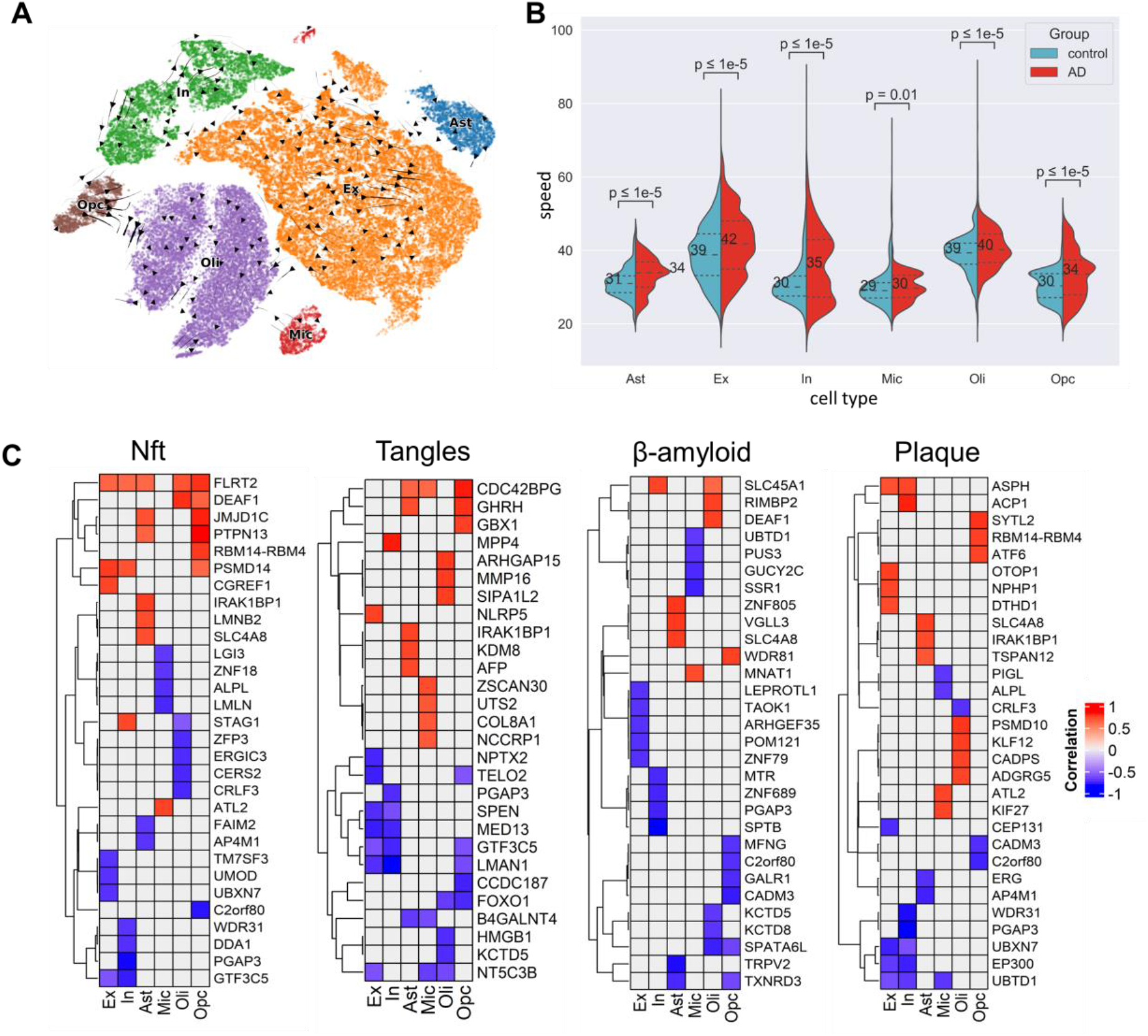
Association of RNA velocity with relevant Alzheimer’s-related neuropathological traits. A) Residual velocity fields between the control and AD cells (z-score > 1.96 or <-1.96). B) Variation of cell speed between the control and AD groups. Median speed overlays plots as numbers (Wilcoxon rank-sum test p-value<0.05) C) Spearman’s correlation of RNA velocity with four AD-neuropathological traits: neuritic plaque count, neuronal neurofibrillary tangle (NFT) counts, overall β-amyloid load (β-amyloid) and PHF tau tangle density (tangles). The confounding effects of age, sex, education, postmortem interval were accounted for via partial correlation. The plots show the top significant genes at FWE-corrected p-value < 0.001.

Lastly, we investigated the association of the RNA velocity differences with four well-known AD pathological traits: neuritic plaque (NP) and neuronal neurofibrillary tangle (NFT) counts based on histochemistry silver stain, and overall β-amyloid load (β-amyloid) and PHF tau tangle density (tangles) based on molecularly specific immunohistochemistry. We calculated Spearman’s correlation between RNA velocity of genes from the 24 AD subjects and the four neuropathological traits while adjusting for the covariates age, sex, education, and postmortem interval. The velocity-phenotype correlations of the top significant genes (FWE-corrected p-value<0.001) are shown in Figure 3C. The genes underlying the different neuropathological are largely cell-specific (Supplementary Figure S3). Only excitatory and inhibitory neurons presented a relatively high overlapping (up to 13%) in significant genes associated with the different AD neuropathological traits. The other cell types substantially differed in gene-specific changes across the phenotypes. Interestingly, we found some AD-relevant genes, including *ADAM10*, associated with tangle burden in excitatory and inhibitory neurons. Some other previously reported AD genes identified include amyloid beta precursor protein binding family members, matrix metallopeptidases, notch, low-density lipoproteins, and protein kinase C’s (Supplementary *Table S3*).

### Cross-study validation of differential RNA velocity

We tested the reproducibility of the observed AD-related differences in RNA velocity in an independent sample. We obtained snRNA-seq data from the dorsolateral prefrontal cortex of another ROSMAP cohort (*Dataset-2*; *Methods, Validation in independent dataset* subsection) comprising 6 cognitively non-impaired individuals with minimal AD pathology and 6 subjects with both pathologic AD and clinical AD dementia (Cain et al. 2020). Following preprocessing, we derived 79,472 cells corresponding to the same six cell types under consideration, and 16,844 transcripts (corresponding to 16,829 unique genes) like in our analysis for the first dataset. Out of the 232 genes with significant RNA velocity differences between the two groups in Dataset-2, 129 (i.e., 56%) overlapped with the genes obtained from the initial analysis of the prefrontal cortex (Dataset-1), including 14 in excitatory neurons, 17 inhibitory neurons, 22 in astrocytes, 34 in microglia, 18 in oligodendrocytes and 24 in oligodendrocytes precursor cells (Figure 4A). The significance of overlap was assessed using Fisher’s exact test (p-value<0.01; odds ratio>1).

**Figure 4.**
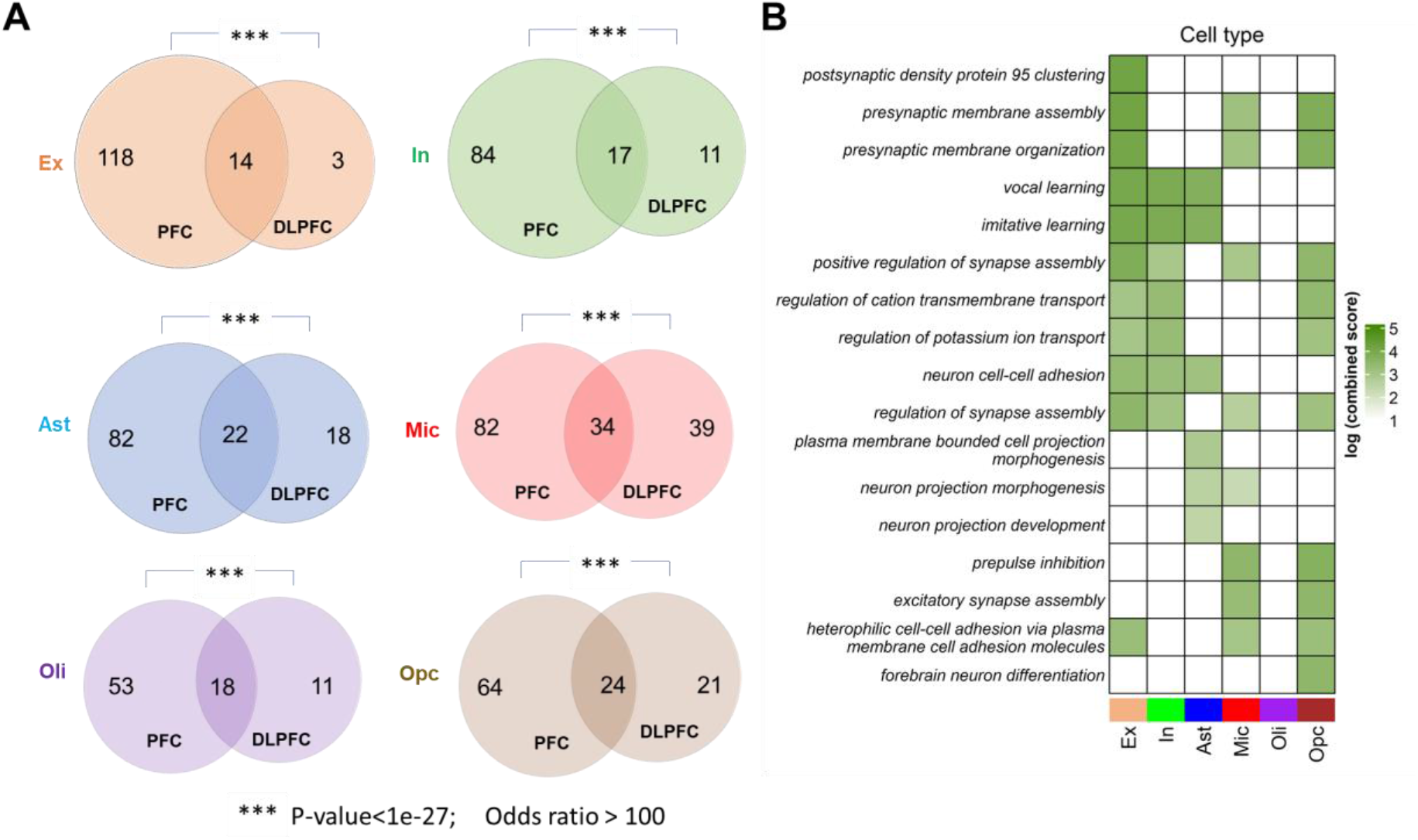
Cross-study validation of RNA velocity differences in neuropathologic AD. A) Venn diagrams for each cell type showing the overlaps between Dataset-1 (prefrontal cortex, PFC) and Dataset-2 (dorsolateral prefrontal cortex, DLPFC) with respect to genes having differential RNA velocity in AD pathology. The sum of all two numbers in any circle represents the number of significant genes in the corresponding dataset. Significance of overlap was estimated with Fisher’s exact test (p-value<0.01; odds ratio>1). B) GO biological processes and their log-transformed EnrichR combined scores for the overlapping gene sets.

We proceeded to query enriched gene ontology terms (GO) of the significant genes common to the two datasets. We used EnrichR tool (Kuleshov et al. 2016; Chen et al. 2013) to uncover the biological processes associated with these overlapping genes. Importantly, we again found that majority of the top biological processes across cell types are associated with neural development and synaptic activities. Overall, these findings support the generalizability of the main RNA velocity differences identified, supporting the robustness of this novel technique for the deep molecular characterization and better understanding of AD pathomechanisms.

## Discussion

Here, we use RNA velocity to characterize, for the first time to our knowledge, the dynamical multicellular processes underlying neuropathological AD progression. Unprecedented advances in scRNA-seq have offered a novel way to overcome the poor spatial resolution of bulk tissue mRNA, while enabling the study of cell type-specific changes in AD and related disorders (Olah et al. 2020; Mathys et al. 2019; Lau et al. 2020). However, differences in RNA expression do not completely capture the evolution of the disease continuum or the progressive vulnerability of cells to neurodegeneration. Using snRNA-seq profiled from postmortem brain samples of the prefrontal cortex and dorsolateral prefrontal cortex in two independent studies, we uncovered highly active genes associated with different levels of a neuropathology. Importantly, the identified cross-validated dynamic genes are associated with a consistent set of molecular functions linked with neurodegeneration. The results support the validity of the novel RNA velocity concept for achieving a complementary molecular characterization of AD and potentially identifying cell-specific disease-modifying genetic targets.

We found accelerated cell dynamics in AD subjects compared to controls, which can explain some of the molecular bases of the early changes occurring in AD. A previous study using induced pluripotent stem cells (iPSC) showed that AD brains undergo accelerated neural differentiation that causes early depletion of neural progenitor pools and reduced cell renewal (Meyer et al. 2019). Further, accelerated cell differentiation may perturb the gene network associated with cell development and synapse organization thereby engendering hyperexcitability and other pathologic cascades. This may as well have implications for the risk of developing dementia as increased cell proliferation of neural progenitor cells in early later and depletion in later life have been linked to APOE deficiency (Yang et al. 2011). Our study suggests that accelerated cell differentiation occurs across different cell types in AD and offers potential areas for experimental validation.

Our analysis revealed that although RNA velocity is closely related to gene expression, the two quantities may capture different pathological processes. The differentially expressed genes associated with AD pathology differed from those associated with varying RNA velocity. The snRNA velocity related genes are principally involved in cell developmental and synaptic programs while the expression-related genes are mainly implicated in ribosomal and mitochondrial activities. In addition to β-amyloid and tau related processes, our analysis of snRNA-vel pointed at other potentially altered functions such as voltage-gated cation channels activity and notch signaling which may have dynamic causal roles in AD development but were not detected by the traditional differential expression. Further, we cross-validated the differential RNA velocity analysis in an independent dataset. The overlapping genes between the two datasets are predominantly implicated in biological processes associated with cell development and synapse organization, implying a recurring theme in our results. A profound nexus exists between cell cycle and synaptic activity in AD, and many AD-associated genes are involved in morphoregulation, i.e., the ordered development and arrangement of cells to form synapse through processes such as cell adhesion, cell differentiation, synaptic membrane assembly, ion channel activity, etc. Besides, results from animal studies showed that certain behaviors simultaneously enhance synaptic plasticity and control accelerated cell cycle, thereby protecting against cell death and neurodegeneration (Arendt 2003).

The RNA velocity metric was designed to capture the dynamic process of cell evolution in the transcriptomic space (La Manno et al. 2018; Bergen et al. 2020). It was originally applied to infer the developmental states of healthy cells but has found further applications in studying cell proliferation in cancer (Couturier et al. 2020; Pan et al. 2020). However, it appears that RNA velocity can also capture dynamic differences associated with severity of AD pathology. Moreover, RNA velocity can be estimated for each cell type at the patient level. Such applications are particularly important for two main reasons. First, prodromal cell changes which occur in AD may be detected before clinical manifestations or the deposition of β-amyloid and tau (Maruszak and Zekanowski 2011; De Strooper and Karran 2016). Second, there are implications for the development of personalized treatment by detecting (and potentially targeting) person-specific contribution of RNA velocity changes to AD neuropathology. We found that most of the biological processes implicated in our study are involved in synapse organization and turnover, a key structural element essential for cognition. Many pathways are also associated with cell developmental processes, the dysfunction of which is linked to neurodegeneration (Joseph et al. 2020). Thus, our results inform potential therapeutic strategies of targeting substrates of synaptic plasticity, including glutamatergic and cholinergic signaling, and applying cell therapy to enhance cell renewal, differentiation, and proliferation.

This study has some limitations. We used single-nucleus RNA sequencing to estimate RNA velocity. Compared to single-cell sequencing, snRNA-seq is more amenable to transcriptomic profiling of postmortem samples because isolated nuclei are intact in frozen tissues (Lake et al. 2017). Moreover, dissociating whole cells from the brain is particularly challenging due to the intensity of the required enzymatic activity, which could interfere with cell type recovery and bias the results of downstream analyses (Habib et al. 2017). RNA velocity was originally formulated for scRNA-seq based on the assumption that the rate of RNA degradation is constant across all cells (La Manno et al. 2018). This assumption is yet to be tested for the rate of nuclear export of spliced RNA into the cytoplasm. Our use of the stochastic model to calculate RNA velocity could limit the bias that may arise from potential differences in the assumptions since degradation is treated as a probabilistic event (Bergen et al. 2020). Importantly, we showed that there is high concordance between RNA velocities from matched nuclear and whole-cell RNA, further supporting the validity of our approach. Recently, RNA velocity was successfully applied to single nuclei data to infer the direction and speed of trophoblast development in the mouse placenta (Marsh and Blelloch 2020).

Changes due to neurodegeneration take years to manifest, but the relatively shorter timescale of mRNA transcriptional dynamics may offer a better resolution to capture the subtle changes that engender pathological cascade. Even though the use many subjects with varied levels of neuropathology allowed us to capture the association between the timescales of transcriptional dynamics and neurodegeneration at the global level, we could not directly ascertain the stability of RNA velocities within a subject over different post-mortem intervals. Nevertheless, we confirmed that the post-mortem sampling intervals between the controls and AD subjects do not differ significantly (Supplementary Figure S4). Future studies can employ multiscale dynamical models of the brain incorporating neuroimaging, gene expression and neuroreceptors can also integrate RNA-vel to better capture the time-resolved complex interactions between those modalities in neurodegenerative progression (Adewale et al. 2021; Khan et al. 2021).

## Conclusions

Our study applies RNA velocity to characterize, for the first time to our knowledge, the dynamical multicellular processes underlying neuropathological AD progression. The generalizability of the results in an independent dataset supports the validity of the novel RNA velocity concept for achieving a complementary molecular characterization of AD, which may be obscured by typical analysis of RNA abundance alone. Specifically, accelerated cell changes were observed in AD. In addition, synaptic and cell developmental processes are implicated in AD neuropathology across cell types. Overall, this initial analysis paves the way for the application of RNA velocity to uncover the dynamical processes in AD and other prevalent neurodegenerative diseases.

## Data Availability

Dataset-1 snRNA-seq and metadata are available at the AMP-AD portal (https://www.synapse.org, IDs syn18485175 and syn3157322). Dataset-2 snRNA-seq and metadata can also be download from the AMP-AD portal (https://www.synapse.org, ID syn16780177). The raw scRNA-seq of Dataset-3 are available on the GEO (https://www.ncbi.nlm.nih.gov/geo/, accession code GSE135618). The data on GEO is freely accessible without registration while data on Synapse are available under controlled use conditions to ensure anonymity of the study participants. Hence, data use agreement and registration are required to access Dataset-1 and Dataset-2. ROSMAP data can be requested at https://www.radc.rush.edu.

## Declarations

### Code Availability

scVelo (v0.2.3) is downloadable as a python package (see https://scvelo.readthedocs.io/installation/). The codes used for the analyses will be available with article publication at https://neuropm-lab.com/other-pipelines.

## Acknowledgements

This project was undertaken thanks in part to the following funding awards to YIM: the Canada Research Chair tier-2, the CIHR Project Grant 2020, the Weston Family Foundation’s Transformation al Research in AD 2020, the Ludmer Centre for Neuroinformatics and Mental Health 2020 award, and the New Investigator start-up grant from McGill University’s Healthy Brains for Healthy Lives Initiative (Canada First Research Excellence Fund). QA is partly supported by Parkinson Canada and Fonds de recherche du Québec – Santé (FRQS) Graduate Partnership Awards, and AFK is partly funded by Parkinson Canada Graduate Student Award. In addition, we used the computational infrastructure supported in part by the Brain Canada Foundation and Health Canada support to the McConnell Brain Imaging Center at the Montreal Neurological Institute. Dataset-1 and Dataset-2 (ROSMAP) were provided by the Rush Alzheimer’s Disease Center, Rush University Medical Center, Chicago. The data collection was supported through funding by NIA grants P30AG10161, P30AG72975, R01AG15819, R01AG17917, R01AG30146, R01AG36836, U01AG32984, U01AG46152, U01AG61356, the Illinois Department of Public Health, and the Translational Genomics Research Institute. Dataset-3 collection was partly supported by National Institutes of Health (NIH) awards F30 AG066418, K08 AG052648, R56 AG057528, K24 AG053435, U54 NS100717, AG023501, AG019724, the Tau Consortium, and the Bluefield Project to Cure FTD.

## Authors’ Contributions

YIM and QA conceived the study. QA and AFK preprocessed the data. QA analyzed the data and wrote the manuscript, with input from YIM and DAB. All authors provided critical feedback and helped shape the research, analysis, and manuscript.

