## Supplementary Figures for "Single-nucleus RNA velocity reveals synaptic and cell-cycle dysregulations missed by gene expression in neuropathologic Alzheimer’s disease"

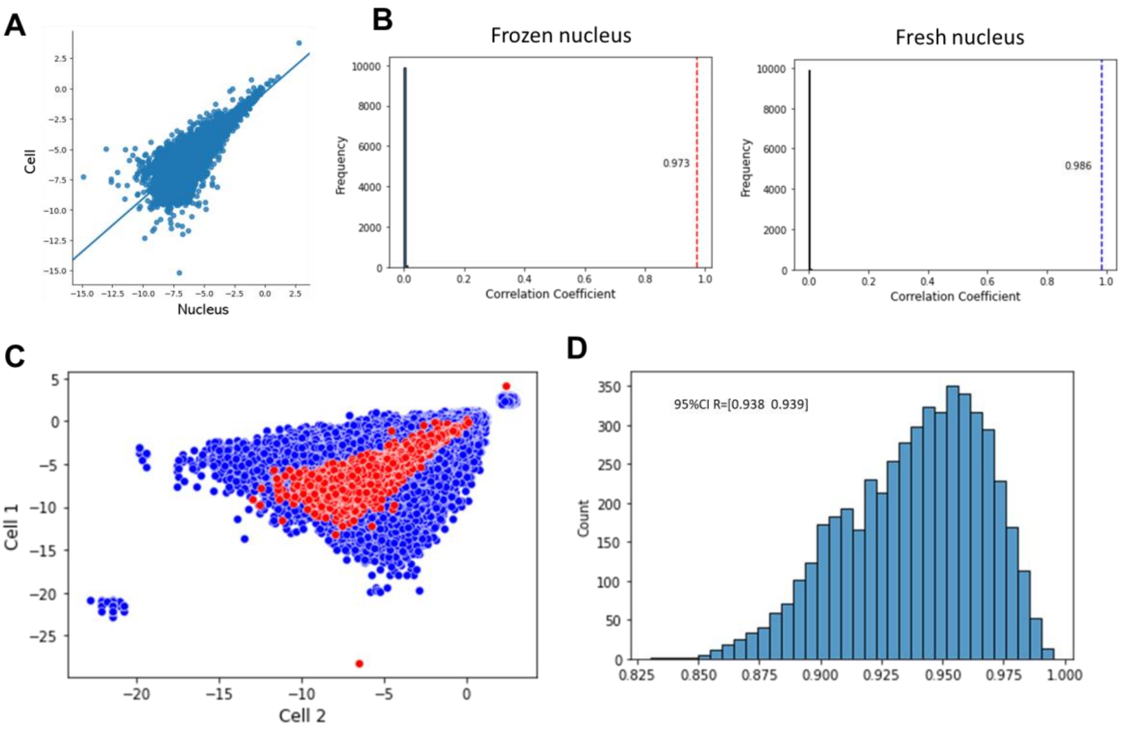


Figure S1 | Validation of the correspondence of RNA velocities single-cell and single-nucleus sequencing dataset of microglia. A) Scatter plot (in log scale) showing the linear concordance between velocities calculated in cells and nuclei for a subject. B) Left: null distribution of Pearson's correlation coefficient between scRNA and frozen snRNA velocities from 5000 permutations. The actual correlation (0.973) is shown in red dotted line. Right: null distribution of Pearson's correlation coefficient between scRNA and fresh snRNA velocities from 5000 permutations. The actual correlation (0.986) is shown in blue dotted line. The second subject (not shown) had correlations of 0.936 and 0.944 in frozen and fresh nuclei, respectively. The high and significant correlations suggest that nuclei RNA can be used to estimate RNA velocities in place of whole-cell RNA. C) Overlay of cell-cell (in blue) and cell-nucleus (in red) RNA velocity correlations. First, a pair of cells is selected at random and the velocities of the genes in the cells are plotted against each other. The random selection is repeated 5000 times to show the variabilities in RNA velocity correlations between single cells. The initial cell-nucleus correlation from A) is then overlayed on the cell-cell correlation plot. The variabilities in cell-nucleus correlation of RNA velocity are observed present in correlations between single cells. D) The correlation distribution between the RNA velocities of any random pair of whole cells. The 95% confidence interval is shown in the plot.

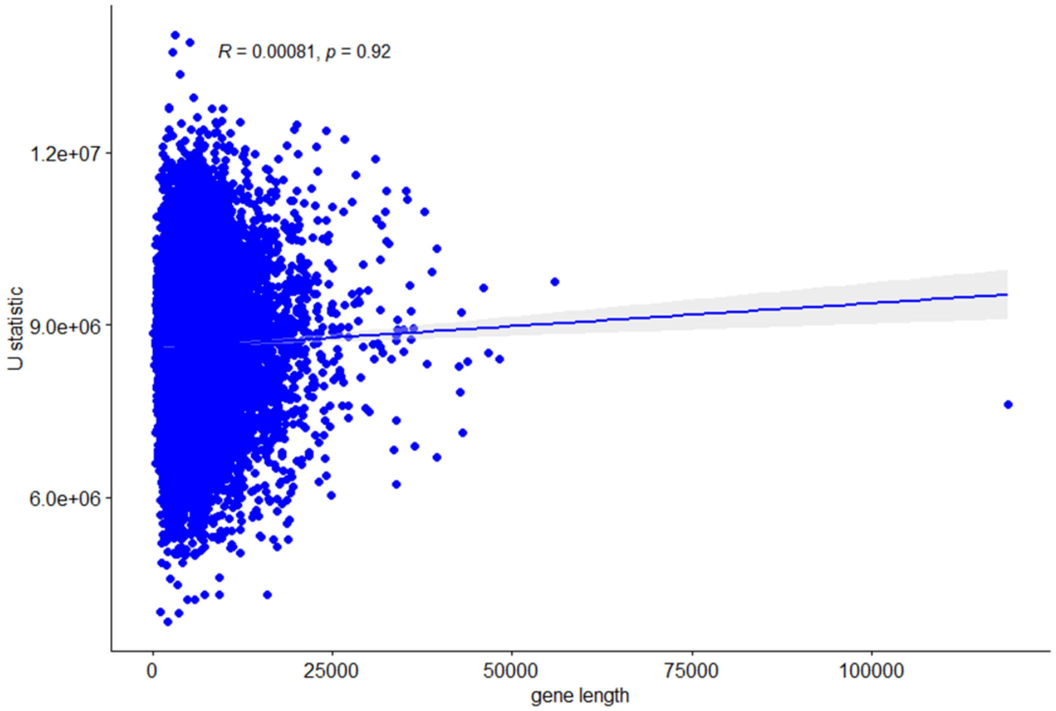


Figure S2 | Correlation between gene length and Wilcoxon rank sum test U-statistic (from differential velocity analysis between AD and control groups) in inhibitory neurons. As there is a tendency for RNA velocities in neurons to be biased towards gene length, we investigated whether the results of differential velocity analyses we performed were influenced by gene length. We found that the correlation between gene length and U-statistic is very minimal and insignificant, supporting that the significant genes identified are not biased by gene length.

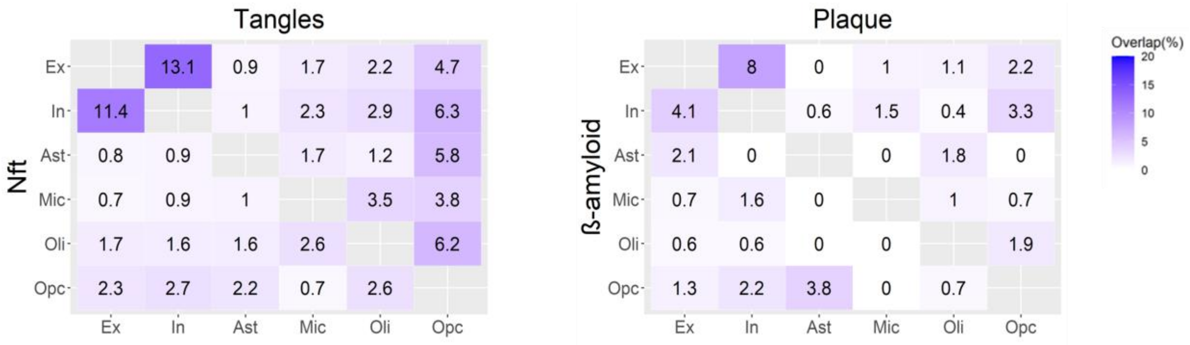


Figure S3 | Percentage overlap of dynamic genes associated with AD pathology in each cell type. The percentage is obtained by expressing the number of genes common to any two cell types as a fraction of the total number of unique genes in the two cell types. Two neuropathological variables are visualized simultaneously on the same plot. Only excitatory and inhibitory neurons presented a relatively high overlapping (up to 13%) in significant genes associated with the different AD neuropathological traits. The other cell types substantially differed in gene-specific changes across the phenotypes.

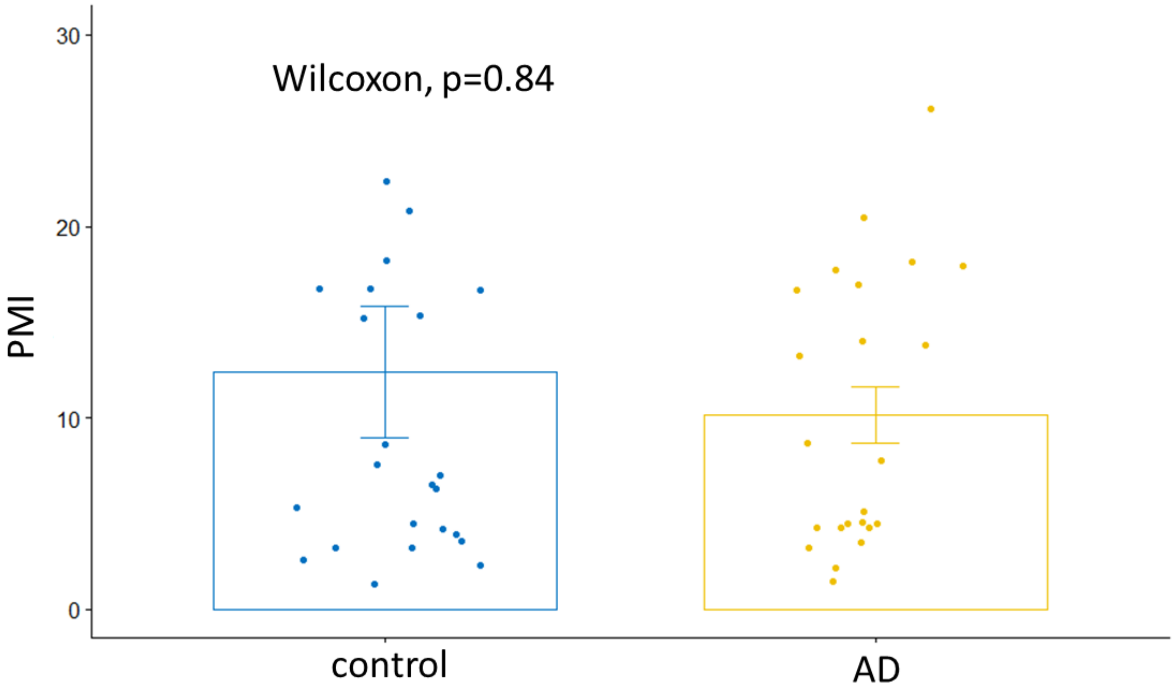


Figure S4 | Test for difference in post-mortem interval between the control and AD groups. Wilcoxon rank-sum test shows that there is no significant difference in the post-mortem sampling intervals of the two groups.
